# Clinical, biochemical and molecular analysis in a cohort of individuals with gyrate atrophy

**DOI:** 10.1101/2023.02.15.23285700

**Authors:** Eleanor Palmer, Karolina M. Stepien, Christopher Campbell, Stephanie Barton, Arunabha Ghosh, Alexander Broomfield, Alison Woodall, Gisela Wilcox, Panagiotis I. Sergouniotis, Graeme C. Black

## Abstract

**Background:** Gyrate atrophy of the choroid and retina is a rare autosomal recessive metabolic disorder caused by biallelic variants in the *OAT* gene, encoding the enzyme ornithine *ƍ*-aminotransferase. Impaired enzymatic activity leads to systemic hyperornithaemia, which in turn underlies progressive chorioretinal degeneration. In this study, we describe the clinical and molecular findings in a cohort of individuals with gyrate atrophy.

**Methods:** Study participants were recruited through a tertiary UK clinical ophthalmic genetic service. All cases had a biochemical and molecular diagnosis of gyrate atrophy. Retrospective phenotypic and biochemical data were collected using electronic healthcare records.

**Results:** 18 affected individuals from 12 families (8 male, 12 female) met the study inclusion criteria. The median age at diagnosis was 8 years (range 10 months – 33 years) and all cases had hyperornithaemia (median: 800 micromoles/L; range: 458-1244 micromoles/L). Common features at presentation included high myopia (10/18) and nyctalopia (5/18). Ophthalmic findings were present in all study participants who were above the age of 6 years. One third of patients had co-existing macular oedema and two thirds developed pre-senile cataracts. Compliance with dietary modifications was suboptimal in most cases. A subset of participants had extraocular features including a trend towards reduced fat-free mass and developmental delay.

**Conclusions:** Our findings highlight the importance of multidisciplinary care in families with gyrate atrophy. Secondary ophthalmic complications such as macular oedema and cataract formation are common. Management of affected individuals remains challenging due to the highly restrictive nature of the recommended diet and the limited evidence-base for current strategies.

## BACKGROUND

Gyrate atrophy of the choroid and retina (also known as ornithine a-aminotransferase deficiency) is an inborn error of metabolism primarily affecting the eye. This rare condition is inherited as an autosomal recessive trait and is associated with biallelic pathogenic variants in the *OAT* gene. *OAT* encodes ornithine aminotransferase, a vitamin B6-dependent enzyme that catalyses the conversion of the urea cycle substrate ornithine to pyrroline 5-carboxylate, which can be subsequently converted into the amino acids glutamate and proline [1,2].

Over 80 pathogenic variants have been described in *OAT* including nonsense, frameshift, splice site and, most commonly, missense variants. These changes lead to a significant reduction in enzymatic activity resulting in the accumulation of ornithine in the plasma (hyperornithaemia), urine and cerebrospinal fluid of affected individuals [3,4]. Notably, high ornithine concentrations are associated with damage to retinal tissue and over time lead to progressive visual loss [5,6,7,8,9,10].

Affected individuals usually present in the first or second decade of life, commonly with night vision problems and/or progressive myopia. [5,6,10,11]. Following this, concentric visual field loss is generally noted during the second decade of life. This slowly progresses to severe loss of vision at around the fourth to fifth decade [5,6,7,8,10,11]. Loss of visual acuity may be secondary to macular oedema and/or photoreceptor cell loss. Early lens opacification is common and most affected individuals require cataract surgery at a relatively young age [12,13,14,15].

The retinal manifestations of gyrate atrophy are highly characteristic and strongly point towards the diagnosis. Fundus examination shows sharply demarcated, scalloped areas of chorioretinal atrophy arising peripherally in the retina. These are initially patchy but gradually enlarge, become confluent and spread centrally towards the macula. Non-ophthalmic manifestations have been described including neonatal hyperammonaemia, intellectual disability, reduced muscle function and bone disorders; some of these findings point to potential long-term consequences of high ornithine levels on other pathways particularly creatine and lysine metabolism [5,6,8,10,11].

There is presently no cure for gyrate atrophy and treatment options remain limited. A low-protein diet with synthetic amino acid supplementation is recommended and has been demonstrated to reduce the rate of retinal degeneration in an animal model [9]. These dietary modifications aim to reduce plasma ornithine levels and to slow disease progression [7,8,9,10,16,17]. However, compliance with diet is often suboptimal, particularly in adolescents and adults, reflecting the difficulties in adjusting to a low protein diet particularly if this is initiated after infancy or early childhood.

Here, we report clinical and genetic findings in 18 individuals with gyrate atrophy. Retinal and systemic manifestations are discussed and the importance of close multi-disciplinary management is highlighted.

## METHODS

### Study participants

Study subjects were retrospectively ascertained through the database of the North West Genomic Laboratory Hub, Manchester, UK. Only families who were diagnosed through the tertiary clinics at Manchester University NHS Foundation Trust and Salford Royal NHS Foundation Trust, Manchester, UK were included. Study participants were referred for genetic testing between 2017 and 2020 and had a homozygous or (at least) two heterozygous pathogenic or likely pathogenic variants in *OAT*.

Ethics committee approval for the study was obtained from the North West Research Ethics Committee (11/NW/0421 and 15/YH/0365) and all investigations were conducted in accordance with the tenets of the Declaration of Helsinki.

### Phenotypic data collection & clinical genetic testing

The clinical notes and/or electronic healthcare record entries were reviewed for each study participant. A full clinical history was obtained and most study subjects were assessed by an ophthalmologist, a metabolic physician and a geneticist. A subset of participants underwent fundus imaging, including widefield retinal imaging, fundus autofluorescence imaging, optical coherence tomography (OCT), and OCT-angiography (OCT-A). The Optos system (Optos PLC, Dunfermline, Scotland, UK) was used to obtain widefield images, the Topcon DRI OCT Triton device (Topcon GB, Newberry, Berkshire, UK) was used to obtain OCT and OCT-A scans, and the Spectralis system (Heidelberg Engineering, Heidelberg, Germany) was used to acquire fundus autofluorescence and OCT images in study participants.

Blood samples were obtained from all probands and DNA was extracted. Multigene panel testing and analysis were subsequently performed at the North West Genomic Laboratory Hub (ISO 15189:2012; UKAS Medical reference 9865) using a previously described approach [18].

## RESULTS

### Clinical and imaging findings

Overall, 18 patients (8 male, 10 female) from 12 families met the study inclusion criteria. This included 15 adults and 3 paediatric patients. Clinical, biochemical and genetic findings in this cohort are shown in Table 1 (See Table 1 in Additional File 1). The majority of study participants (n=17) had south-east Asian ancestries.

The median age at diagnosis was 8 years (range: 10 months – 33 years). All affected individuals had plasma ornithine levels above the normal range at the time of diagnosis (range: 458 – 1244 µmol/L) (reference ranges: 40-150 µmol/L, provided by the Willink Metabolic Laboratory, which was responsible for analysis of all samples referenced in this study). Common features at presentation included myopia (10/18 cases) and night vision problems (5/18 cases). Ophthalmic findings were present in all study participants who were above the age of 6 years. The median age when chorioretinal changes were first clinically documented was 13 years (range: 4– 33 years). The median visual acuity at the most recent ophthalmic assessment was 0.36 LogMAR units (range: 0.0–0.7 LogMAR). One third of study participants (6/18 cases) had macular oedema which was managed by carbonic anhydrase inhibitors, including oral acetazolamide in most (5/6) cases. Notably, 4 of the 6 patients affected by macular oedema were also noted to develop pre-senile cataracts. Overall, 12 of the 18 study participants (66.6%) developed pre-senile cataracts in early adulthood; 7 of these cases underwent cataract surgery with the median age of intervention for this subset being 27.5 years (range: 22–36 years). Five affected individuals were clinically documented as sight impaired registered, though limited data were available to confirm registration for most patients. Other ophthalmic features included epiretinal membrane (n=3), floaters (n=2), retinoschisis (n=1), optic nerve drusen (n=1), chronic anterior uveitis (n= 2), amblyopia (n=1) and exophoria (n=1).

The median body mass index (BMI) for the adult subset of the cohort (15/18) was 23.4 kg/m^2^. Six participants were found to have learning difficulties (33%) and 13 study subjects (72%) had other extraocular features including those listed in Table 2. (See Table 2 in Additional File 2).

Retinal imaging data were reviewed. Five study participants were not imaged in the 5 years preceding data collection, one of whom was an infant. Due to the ongoing SARS-CoV-2 pandemic, the recent provision of imaging was found to be reduced. Over the past 5 years, the group of patients who were imaged received some form of imaging a median of 8 times (mean: 9.5 times, range: once to 22 times). With regards to OCT imaging, individuals in this subset had imaging a median value of four times in the past 5 years (mean: 4.1, range: 7 months to >60 months). OCT-A imaging was not frequently performed in this cohort as only two patients were imaged using this modality in the past 5 years. Fundus autofluorescence and widefield retinal images were obtained on average once every 12 months (median 12: range 5 months to >60 months).

### Biochemical findings

Biochemical data are presented in Table 3 (See Table 3 in Additional File 3). The median plasma ornithine level at diagnosis was 800 µmol/L (range: 458-1244 µmol/L). As stated previously the median age at time of diagnosis was 8 years (range: 10 months – 33 years). The cohort was divided into those participants diagnosed by the age of 5 years or less (n =6), and those diagnosed at age 6 years or older (n=12). The 6 individuals who were diagnosed at age 5 years or younger had a median plasma ornithine of 719 µmol/L (mean: 753, range: 458-1232). The median plasma ornithine over the last 5 years for this subset was 581 µmol/L (mean: 626, range: 392-879), representing a 19% decrease. Of the 12 patients diagnosed above age 6 years, 9 had complete data for plasma ornithine levels at the time of diagnosis and in the last 5 years prior to data collection. Those diagnosed after the age of 6 years had a median plasma ornithine of 800 µmol/L at diagnosis (mean: 844, range 576-1120) and 716 µmol/L (mean: 787, range 528-1342), representing an 11% decrease.

At the time of diagnosis, all study participants were advised to follow a low protein diet with additional essential amino acid supplementation in an attempt to lower plasma ornithine. For adults, the recommended target for natural protein, depending on the individual, was between 0.6 and 0.8 g/kg/day. If protein intake was reported as less than 0.8g/kg/day, essential amino acid supplementation was recommended with the aim of achieving a total protein intake of 1g/kg/day. Other supplements included pyridoxine (in case of pyridoxine responsive OAT deficiency), lysine (to competitively reduce renal tubular reabsorption of ornithine), and creatine (to correct the creatine deficiency resulting from inhibition of synthesis by elevated plasma ornithine) [15]. Over the past 5 years, the median plasma ornithine for the whole cohort was 678.5 µmol/L (range: 392-1342 µmol/L), representing an overall percentage decrease of 15% compared to the plasma ornithine at the time of diagnosis.

Information on dietary compliance was collected for 15 patients within the cohort. Six patients did not restrict dietary protein at all (Cases 1, 2, 5, 11, 12, and 15). This subset had a median plasma ornithine at time of diagnosis and over the past 5 years of 771 µmol/L (mean: 852 µmol/L) and 717 µmol/L (mean: 841 µmol/L) respectively, representing a 7% decrease. The median visual acuity at the time of last ophthalmic examination for participants who did not restrict protein intake was 0.45 logMAR (range: 0.31 – 0.7 logMAR) at a median patient age of 26 years. For those individuals who did restrict protein intake and for whom complete data was available (n=8), the median plasma ornithine at time of diagnosis, and over the past 5 years, was 764.5 µmol/L (mean: 761 µmol/L) and 597 µmol/L (mean: 668 µmol/L) respectively, representing a 22% decrease. For this subset who did limit dietary protein, the median visual acuity recorded at last ophthalmic examination was 0.34 logMAR (range, 0.0 – 0.92 logMAR) at a median age of 21 years.

The most frequently utilised amino acid supplement within the cohort (n=7) was Essential Amino Acids (EAA) (Vitaflo International Ltd, Liverpool, UK) at a dosage of between 3-4 sachets per day on average. Cases 10, 11, 12, 13 took a vitamin D supplement in addition to their current daily supplement regime (2000 units daily). Cases 1 and 10 took an additional folic acid supplement. Case 10 also took a ferrous sulphate supplement. Cases 1, 5, 13 and 15 took additional creatinine monohydrate peptide supplementation. The median plasma lysine over the past 5 years in the rest of the cohort was 124 µmol/L (range 72-247 µmol/L). 6 out of 15 patients had plasma lysine <100 µmol/L (normal range 100-260 µmol/L) and 7 patients required supplementation (lysine 4 g/day). Two study participants (cases 2 and 4) saw an increase in plasma ornithine from the time at diagnosis compared with the overall average over the past 5 years; this was probably related to their poor dietetic compliance (see Table 3 in Additional File 3).

### Genetic findings

Overall, 14 different likely disease-causing DNA variants in *OAT* were detected. Five of these changes have not been previously reported (See Table 4 in Additional File 4). The variants identified in the cohort include missense (n=7), nonsense (n=3), frameshift (n=2) and splicing (n=2) variants.

Of the previously unreported variants identified in this study, *OAT* c.1009dupC p.(Leu337ArgfsTer2) is a pathogenic frameshift change in exon 8 of the gene, which is predicted to elicit nonsense-mediated decay. This variant was identified in a heterozygous state with another novel heterozygous missense variant, c.1208T>C p.(Leu403Pro), in case 17. This missense change affects a highly conserved leucine residue and *in silico* evidence predicts that a proline substitution at this position will have a damaging effect. Notably, this is the only participant in the cohort who has European ancestries. The phase of the two variants has not been established for this case, however the same genotype has been detected in an unrelated proband, in whom compound heterozygosity was proven by parental testing (unpublished data).

Another previously unreported variant is c.520+1G>A. This change affects the canonical splice donor site in intron 4 of the *OAT* gene and is predicted to cause in-frame skipping of exon 4. This likely pathogenic variant is absent from the “controls/biobanks” subset of the Genome Aggregation Database (gnomAD v.2.1.1) dataset and is present in the homozygous state in the three affected members of a family in this cohort. Another novel variant, *OAT* c.648G>C p.?, was present in the homozygous state in case 1 (Table 4). This change is also predicted to cause aberrant splicing due to the substitution at the last nucleotide position of exon 5.

The final novel change was a missense variant, c.941T>G p.(Ile314Ser), which was identified in a heterozygous state in case 15 in addition to a heterozygous pathogenic c.748C>T p.(Arg250Ter) nonsense variant; parental testing has not been performed to confirm that these variants are *in trans*. The isoleucine residue at position 314 is highly conserved and multiple lines of computational evidence predict that the substitution of a serine residue at this codon will have a deleterious effect on the *OAT* protein function.

## DISCUSSION

This report describes observational data collated for a cohort of individuals with gyrate atrophy. Hyperornithaemia and changes in the *OAT* gene were present in all study participants. Diagnosis of gyrate atrophy is frequently achieved surprisingly late. In this cohort, the median age at diagnosis was 8 years. This is comparatively earlier than that described in a systematic review [15] where a median age of diagnosis of 13 years was reported amongst 47 patients. Amongst the patients described here, some remained asymptomatic until adolescence and were diagnosed either on a routine eye assessment or through family screening. Notably, most patients were of Southeast Asian origin suggesting an increased incidence within this population; most also had a history of consanguinity.

A multidisciplinary approach is key for the management of individuals with gyrate atrophy. Ophthalmic imaging is an important investigation in this patient group as it can help monitor disease progression. In this cohort, there was a high degree of variability in terms of the frequency of imaging studies. This may be due to several underlying factors including patient disengagement, the impact of the SARS-CoV-2 pandemic or barriers in communication between service departments. No current standardised guidelines exist for disease monitoring using imaging and different centres utilise varying imaging protocols (utilising modalities such as fundus autofluorescence, fluorescein angiography [19] and OCT in addition to fundoscopy). Notably, a large proportion of study participants (78%) experienced macular oedema and/or pre-senile cataracts in addition to core manifestations of nyctalopia and peripheral vision deficits. Macular oedema is a known complication of gyrate atrophy [19,20], and supportive therapy including administration of topical or oral carbonic anhydrase inhibitors, topical non-steroidal anti-inflammatory drops, intravitreal corticosteroids or intravitreal anti-vascular endothelial growth factor agents has been reported to have a favourable short-term impact on retinal structure [15,20]. However, large-scale studies are lacking and the effect of these treatments on visual function remains uncertain. Pre-senile posterior subcapsular cataracts are common in individuals with gyrate atrophy and early lens surgery is often required. It is noted that among the subgroup of study participants that required cataract surgery (n=6), only 50% of cases had macular oedema; this highlights that the link between prolonged hyperornithaemia and distinct ophthalmic complications is likely to be complex. Interestingly, the visual acuity of the study participants who restricted dietary protein was better on average than that of those who did not observe any protein limitation. Although this difference may be due to confounding factors or change, this observation is in keeping with reported evidence that dietary manipulation may correlate with better visual outcomes [7,8,9,10,15,16,17]. Despite this, progressive visual loss was observed in all patients, highlighting that current management strategies remain suboptimal.

A variety of non-ophthalmologic manifestations was noted in this cohort. The most common finding, observed in one third of patients, was developmental/cognitive impairment. Although learning difficulties have been reported in the literature as a frequently observed patient finding in association with systemic hyperornithaemia and secondary creatine deficiency [1,2], our current understanding of the expression of gyrate atrophy related neurocognitive phenotypes is limited. There is similarly lacking evidence pertaining to any correlation between the severity of these manifestations and the levels of raised plasma ornithine. Early studies by Kaiser-Kupfer and colleagues [19] reported abnormalities in electroencephalographic testing in individuals with gyrate atrophy, although these did not appear to correlate with any clinical neurocognitive dysfunction. As above stated, the majority of patients in this cohort were born to consanguineous parents, raising the possibility of other genetic factors affecting intellectual outcome.

Similarly, fat free mass biopsy in individuals with gyrate atrophy may reveal abnormalities due to the metabolic effects of raised ornithine and secondary creatine deficiency [1,15,21]. However, not all patients with gyrate atrophy exhibit muscular dysfunction [15]. When assessed in clinic, 9 of 11 patients in this cohort had body composition analysis by SECA mBCA Multi-frequency Bioelectrical Impedence Analysis, which indicated a reduced fat free mass. These patients commenced therapy with creatine monohydrate (1.5 mg four times per day) in an attempt to increase fat free mass and muscle strength [22]. Reports of reduced fat free and/or muscle mass in patients with gyrate atrophy need comparison with findings of reduced fat-free mass, by gold standard methodology, in young adults with a range of inherited disorders of protein metabolism as protein restriction alone may be contributory [23].

Given the rarity of the condition, robust data on the management and long-term outcomes in gyrate atrophy are limited. Further, as with other rare genetic disorders, obtaining data on which evidential management can be based remains challenging. Most of the evidence-base for current management strategies involves studies from the 1980s and 1990s [7,8,9,10,15,17]. No previous randomised control trials or large cohort studies have been described in the biomedical literature [15]. Looking forward, multicentre, collaborative research is required to evaluate available imaging modalities and to develop diagnostic and monitoring recommendations (guidelines) for the long-term surveillance, intervention and treatment outcomes. Furthermore, there is a clinical need for better referral diagnostic pathways to ensure that early dietary modification is offered to newly confirmed cases.

Our findings support the proposal that dietary manipulation may reduce plasma ornithine [16,17] and that this in turn may confer positive outcomes in terms of limiting vision loss. Furthermore, those individuals diagnosed at an earlier age had lower baseline plasma ornithine levels compared to those diagnosed later. What is often understated however is that the recommended protein restriction imposes a strict and life-long limitation and that patient compliance has been historically poor (less than 70% in this study) due to the poor palatability of dietetic products [17]. Studies investigating the quality-of-life impact of current management strategies for gyrate atrophy are lacking and a balanced assessment of the patient impact of such approaches would help guide future management. In addition to patient challenges with dietary restriction, frequent hospital attendance for appointments with ophthalmologists, metabolic medicine physicians and dieticians can be problematic. Notably, the fact that most affected individuals do not perceive the clinical impact of reducing protein intake and taking supplements/vitamins, can lead to poor engagement and poor compliance. A multidisciplinary approach with a formalised retinal imaging programme is recommended and additional support when transitioning from paediatric to adult services is required. The earlier the low protein diet is introduced, the better compliance in adolescence and adulthood is expected to be. It is noted that participants with learning difficulties who were fully dependent on their parents/carers seemed to follow stricter diet and their compliance with amino acid products was found to be much better.

In conclusion, this report expands knowledge of gyrate atrophy. The restriction of dietary protein reduces plasma ornithine but compliance with treatment can be challenging and a personalised approach is required. Despite the maxim of gyrate atrophy being a “treatable” condition, management remains suboptimal due to poor patient compliance and lack of an effective treatment. Future research into therapies for gyrate atrophy, including novel gene-based interventions such as those utilised in other inherited retinal disorders, would be of significant interest and has the potential to revolutionise clinical management.

## Data Availability

All data generated or analysed during this study are included in this published article.

## Acknowledgement

The authors would like to thank Stuart Ingram and Jingshu Liu for their contribution to the data collection

## ADDITIONAL FILES

**Additional file 1** - Table 1: Molecular diagnosis, clinical and biochemical findings in a cohort of 18 patients with gyrate atrophy

**Additional file 2** - Table 2: Comorbidities observed in 13 individuals with gyrate atrophy. [Body composition measured in an outpatient clinic setting using a SECA mBCA Bio-Impedance Analysis Machine and completed as per manufacturer’s instructions.]

**Additional file 3** - Table 3: Biochemical and visual outcomes at most recent clinic visit using current management strategies

**Additional file 4 -** Table 4: Genetic Variants and *in silico* analysis

