## Supplemental Tables 1-4 for "Clinical, biochemical and molecular analysis in a cohort of individuals with gyrate atrophy"

**Table 1: Molecular diagnosis, clinical and biochemical findings in a cohort of 18 patients with gyrate atrophy**

| **Patient No.** | **Gender** | **Age Range at Last Ophthalmic Examination (years)** | **Visual Acuity**  **(LogMAR)** | | **Presenting Ophthalmic Symptom(s)**  **(Age at Diagnosis [years])** | **Plasma Ornithine Levels at Diagnosis (μmol/L)** | **Average Plasma Ornithine in Last 5 Years**  **(μmol/L)** | **Average Plasma Lysine in Last 5 Years (μmol/L)**  **[NR: 100-160 μmol/L]** | **Genetic Diagnosis, Pathogenic/Likely Pathogenic Variant** |
| --- | --- | --- | --- | --- | --- | --- | --- | --- | --- |
|  |  |  | **Right eye** | **Left eye** |  |  |  |  |  |
| 1 | M | 31-35 | 0.4 | 1.0 | Increasing myopia (14) | 1120 | 1067 | 87 | c.520+1G>A (homozygous) |
| 2 | M | 21-25 | 0.68 | 0.54 | Increasing myopia (9) | 1018 | 1342 | 73 | c.520+1G>A (homozygous) |
| 3 | F | 26-30 | 0.22 | 0.24 | Increasing myopia (11) | 1007 | 863 | 72 | c.520+1G>A (homozygous) |
| 4 | M | 15-20 | 0.32 | 0.44 | Asymptomatic (2) | 458 | 868 | 84 | p.(Arg398Ter) (homozygous) |
| 5 | F | 26-30 | 0.42 | 0.3 | Asymptomatic (5) | 695 | 564 | 87 | p.(Arg398Ter) (homozygous) |
| 6 | F | 21-25 | 0.34 | 0.34 | Increasing myopia (6) | 1244 | N/A | N/A | p.(Arg398Ter) (homozygous) |
| 7 | F | 5-14 | 0.1 | 0.1 | Increasing myopia (8) | 576 | 528 | 150 | p.(Pro241Leu) (homozygous) |
| 8 | M | 5-14 | 0.38 | 0.3 | Asymptomatic (4) | 754 | 597 | 130 | p.(Pro241Leu) (homozygous) |
| 9 | F | 21-25 | 0.5 | 0.64 | Nyctalopia, Reduced peripheral vision (8) | 775 | 640 | 106 | p.(Pro241Leu) (homozygous) |
| 10 | F | 31-35 | 0.94 | 0.9 | Increasing myopia, Nyctalopia, Reduced peripheral vision (7) | 826 | 572 | 76 | p.(Pro241Leu) (homozygous) |
| 11 | F | 26-30 | 0.5 | 0.6 | Increasing myopia, Nyctalopia, Cataracts (26) | 800 | 718 | 204 | p.(Pro241Leu) (homozygous) |
| 12 | F | 26-30 | 0.3 | 0.4 | Increasing myopia, Nyctalopia (14) | 734 | 641 | 103 | p.(Gly51Asp) (homozygous) |
| 13 | M | 21-25 | 0.16 | 0.3 | Asymptomatic (0.83) | 458 | 392 | 169 | p.(Arg398Ter)  (homozygous) |
| 14 | M | 0-4 | 0.0 | 0.0 | Asymptomatic (1) | 1232 | 879 | 195 | p.(Pro241Leu); (heterozygous) p.(Gly353Asp) (heterozygous) |
| 15 | F | 21-25 | 0.22 | 0.4 | Increasing myopia (14) | 742 | 716 | 108 | p.(Arg250Ter);  (heterozygous)  p.(Ile314Ser)  (heterozygous) |
| 16 | F | 41-45 | 0.0 | -0.1 | Increasing myopia, Nyctalopia, Cataracts (33) | 917 | N/A | N/A | p.(Tyr209Ter); (heterozygous) p.(Pro417Leu) (heterozygous) |
| 17 | M | 15-20 | 0.22 | 0.5 | Increasing myopia, Floaters (14) | N/A | 458 | 247 | p.(Leu403Pro); (heterozygous) (Leu337ArgfsTer2) (heterozygous) |
| 18 | M | 26-30 | 0.48 | 0.6 | Asymptomatic (5) | 920 | N/A | N/A | p.(Pro300LeufsTer13) (homozygous) |

**NB: Two patients for whom genetic data only were available have been excluded from the above table and information regarding their genetic diagnosis is included in Table 4.**

**Table 2: Comorbidities observed in 13 individuals with gyrate atrophy**

| **Organ System** | **Comorbidity** | **Number of patients affected** |
| --- | --- | --- |
| Psychological/Neurological | Developmental delay | 6 |
|  | Depression | 2 |
|  | Psychosis | 1 |
|  | Anorexia nervosa | 1 |
|  | Dyslexia | 1 |
|  | Dyspraxia | 1 |
|  | Spastic dysplegia | 1 |
|  | Reduced fat-free mass | 9 |
| Haematological | Anaemia (unspecified) | 1 |
|  | Congenital dyserythropoeitic anaemia | 1 |
|  | Red cell degeneration | 1 |
| Respiratory | Asthma | 2 |
|  | Tuberculosis | 1 |
|  | Vocal cord nodules | 1 |
| Cardiovascular | Congenital heart disease | 1 |
| Endocrinological | Precocious puberty | 1 |
|  | Diabetes insipidus | 1 |
| Urological | Dysfunctional bladder syndrome | 1 |

**Table 3: Biochemical and visual outcomes at most recent clinic visit using current management strategies**

| **Patient no.** | **Compliance with current treatment (Y/N/P)§** | **Protein restriction (daily intake [g/kg] excluding EAA)** | **Daily Lysine Supplementation (g)** | **Current additional amino acid supplement regime with total daily dosage (where available)** | **Plasma ornithine at diagnosis (NR: 40-150 μmol/L)** | **Average plasma ornithine in last 5 years (μmol/L)** | **Average plasma lysine in last 5 years**  **(NR: 100-160 μmol/L)** | **Average visual acuity at last ophthalmic examination (logMAR)** |
| --- | --- | --- | --- | --- | --- | --- | --- | --- |
| 1 | P | No restriction | 5 | None | 1120 | 1067 | 87 | 0.7 |
| 2 | N | No restriction | 0 | None | 1018 | 1342 | 73 | 0.61 |
| 3 | P | 0.5-0.6 | 0 | None | 1007 | 863 | 72 | 0.23 |
| 4 | Y | 1 | 4 | EAA (3-4 sachets) | 458 | 868 | 84 | 0.38 |
| 5 | P | No restriction | 4 | EAA (3-4 sachets) | 695 | 564 | 87 | 0.36 |
| 7 | Y | 0.5-0.8 | 0 | EAA (4 sachets)  UCD Amino 5 (1 sachet)  Dialamine (100g) | 576 | 528 | 150 | 0.1 |
| 8 | Y | 0.2-0.3 | 0 | EAA (4 sachets) | 754 | 597 | 130 | 0.34 |
| 9 | Y | 1 | 4 | None | 775 | 640 | 106 | 0.57 |
| 10 | Y | 0.7-1 | 10 | None | 826 | 572 | 76 | 0.92 |
| 11 | P | No restriction |  | EAA  Pyridoxine (300mg) | 800 | 718 | 204 | 0.55 |
| 12 | P | No restriction | 10 | EAA (4 sachets) | 734 | 641 | 103 | 0.35 |
| 13 | Y | 0.15 | 0 | Dialamine (22 scoops)  Phlexyvits  Zinc biotin | 458 | 392 | 169 | 0.23 |
| 14 | Y | 0.4 | 0 | EAA (3 sachets)  Docomega | 1232 | 879 | 195 | 0.0 |
| 15 | Y | No restriction | 4 | None | 742 | 716 | 108 | 0.31 |
| 17 | Y | 0.5-0.6 | 0 | Dialamine (100g)  Fruityvit | **+** | 458 | 247 | 0.36 |

**Cases 7, 8 and 14 are paediatric patients; cases 6, 16 and 18 had no available recorded data on ornithine or lysine levels.**

**§ Y/P/N: Yes = compliant with both protein restriction, Partial = compliant with supplementation but not protein restriction, N = not compliant. Compliance was determined by patient metabolic medicine/dietician appointments**

**+ Data not available**

**Table 4: Genetic Variants and *in silico* analysis**

| **HGVS description (NM_000274.4)** | **REVEL score** | **GnomAD (v2.1.1)**  Number of non-ref alleles / total number of alleles | **Reference**  (Clinvar & HGMD) |
| --- | --- | --- | --- |
| c.520+1G>A | n/a | Absent | Not reported on HGMD |
| c.461G>A p.(Arg154His) | 0.929 | 2/251370 | Ghosh et al. 2017 |
| c.1192C>T p.(Arg398Ter) | n/a | 2/250954 | Michaud et al. 1995 |
| c.722C>T p.(Pro241Leu) | 0.8999 | 9/251486 | Brody et al. 1992 |
| c.152G>A p.(Gly51Asp) | 0.875 | 3/251488 | Sergouniotis et al. 2012 |
| c.648G>C p.? | n/a | Absent | Not reported on HGMD |
| c.899delC p.(Pro300LeufsTer13) | n/a | Absent | Patel et al. 2018 |
| c.1058 G>A p.(Gly353Asp) | 0.9219 | 10/250560 | Brody et al. 1992 |
| c.748C>T p.(Arg250Ter) | n/a | 2/251456 | Sergouniotis et al. 2012 |
| c.941T>G p.(Ile314Ser) | 0.9169 | Absent | Not reported on HGMD |
| c.627T>A p.(Tyr209Ter) | n/a | 6/282864 | Mashima et al. 1992 |
| c.1250C>T p.(Pro417Leu) | 0.8889 | 8/282378 | Brody et al. 1992 |
| c.1208T>C p.(Leu403Pro) | 0.9449 | 2/282564 | Not reported on HGMD |
| c.1009dup p.(Leu337ArgfsTer2) | n/a | Absent | Not reported on HGMD |

Ghosh et al. <https://pubmed.ncbi.nlm.nih.gov/28468868/>

Michaud et al. <https://pubmed.ncbi.nlm.nih.gov/1612597/>

Brody et al. <https://pubmed.ncbi.nlm.nih.gov/1737786/>

Sergouniotis et al. <https://pubmed.ncbi.nlm.nih.gov/22182799/>

Patel et al. <https://pubmed.ncbi.nlm.nih.gov/30054919/>

Mashima et al. <https://pubmed.ncbi.nlm.nih.gov/1609808/>
